# An essential medicines list in Ireland: A qualitative interview study of interest-holders

**DOI:** 10.1101/2025.08.26.25334433

**Authors:** James Larkin, Matthew Preteroti, Logan T. Murry, Michelle Flood, Barbara Clyne, Sara Burke, Tom Fahey, Nav Persaud, Frank Moriarty

**Affiliations:** School of Pharmacy and Biomolecular Sciences, RCSI University of Medicine and Health Sciences, Dublin, Ireland; Department of General Practice, RCSI University of Medicine and Health Sciences, Dublin, Ireland; Department of Epidemiology and Public Health, School of Population Health, RCSI University of Medicine and Health Sciences, Dublin, Ireland; Centre for Health Policy and Management, School of Medicine, Trinity College Dublin, Dublin, Ireland; Department of Family and Community Medicine, University of Toronto, Toronto, Ontario, Canada; MAP Centre for Urban Health Solutions, St Michael’s Hospital, Toronto, Ontario, Canada

## Abstract

**Background:** The World Health Organization recommends each country develop a national essential medicines list (NEML), prioritising a core medicines set aligned with national health needs. Policy in Ireland partly focusses on prescribing costs and quality, which creates opportunities for NEML development. We aimed to explore interest-holders’ perspectives on an Irish NEML.

**Methods:** We applied a descriptive qualitative methodology. Using purposive and snowball sampling, we recruited interest-holders from Irish bodies/groups with roles in shaping Ireland’s medicines policy/use with potential for involvement in a NEML. Semi-structured interviews were conducted and analysis involved Braun and Clarke’s six-stage approach to thematic analysis.

**Results:** Thirteen participants were interviewed and three themes were generated: 1) the NEML’s purpose, 2) NEML barriers and facilitators and 3) development and implementation processes.

For participants, an NEML’s purpose is meeting the population’s priority needs. Participants also outlined roles in ensuring adequate supplies and as a national formulary. Views differed on whether an NEML should involve reduced costs to patients for access. Participants proposed that the national government health department, the state body who run the health service (HSE), and/or the medicines regulator (HPRA) should be responsible for an NEML.

**Conclusions:** Participants perceived an NEML in Ireland as beneficial and aligning with the WHO’s vision: medicines that effectively and safely treat the priority healthcare needs of the population. Future work should explore the patients’ and the public’s perspectives on an NEML. Other countries’ NEMLs offer exemplars to inform Ireland’s approach.

**Research in Context:** *What is already known about the topic?:* According to The World Health Organization, "each country has the direct responsibility of evaluating and adopting a list of essential drugs, according to its own [health] policy." These national essential medicines lists (NEMLs) can be used to standardising medications used across healthcare institutions. This can have benefits for procurement costs, patient safety and preventing shortages.

*What does this study add to the literature?:* We interviewed 13 medicines policy interest-holders about an Irish NEML. Participants envisioned an NEML as meeting the population’s priority needs. They also outlined roles for an NEML in ensuring supplies and as a national formulary. Barriers (e.g. achieving consensus) and facilitators (e.g. buy-in) to developing and implementing an NEML were outlined. Participants said transparent development processes, the use of financial incentives and a communications strategy, could help overcome barriers. Participants proposed that the national government health department, the state body who run the health service (HSE), and/or the medicines regulator (HPRA) should be responsible for an NEML.

*What are the policy implications?:* An NEML could fit into Ireland’s ongoing health reform policy: Sláintecare. Participants’ suggested strategies for overcome NEML barriers are backed-up by findings in other high-income countries (e.g. Sweden and Canada) where transparency, financial incentives and a communications strategy contributed to NEML success.

## 1. Introduction

The World Health Organization (WHO) has stated that "each country has the direct responsibility of evaluating and adopting a list of essential drugs, according to its own policy in the field of health" [1]. Several countries have used the WHO Model List of Essential Medicines to guide the development of national essential medicines lists (NEMLs), which prioritise a core set of medicines based on each country’s health needs and guide medicine selection and appropriate use [2, 3]. In the context of national lists, ‘essential’ indicates that some medicines may be considered higher priority due to population-specific health needs or specific clinical or prescribing recommendations, with calls for increased access to these medicines [4, 5].

Benefits associated with the implementation of NEMLs, include standardising medications used across healthcare institutions and opportunities for bulk buying, with cost-containment effects as medication procurement costs can be reduced [6, 7]. A study evaluating the implementation of an NEML in India identified that this contributed to savings of 30% on annual government medicine costs [7]. NEMLs can also minimise the frequency and magnitude of medication shortages, encouraging supply of priority medications [4, 8, 9]. Therefore, along with possible cost-containment benefits, NEMLs can positively impact patient safety. Noting the recommendations by the WHO and the positive effects of NEMLs, Ireland can potentially benefit from an NEML.

With ongoing policy and reform initiatives in Ireland, like the Preferred Drugs Initiative [10], focused on improving prescribing quality and decreasing costs, there is potential for an NEML specific to the needs of Ireland. It is important to explore the perspectives of key health interest-holders in the creation of NEMLs [11, 12]. Therefore, this study aims to explore the perspectives of interest-holders on the purposes, benefits, processes, and challenges surrounding the development and implementation of an Irish NEML.

## 1. Methods

### 2.1 Study design

This study applied a descriptive qualitative methodology [13] to provide a comprehensive understanding of the perspectives of interest-holders in Ireland about an Irish NEML. This study was reported according to the Consolidated Criteria for Reporting Qualitative Research reporting guidelines (eTable 1) [14]. The study was approved by the RCSI Human Research Ethics Committee (REC202003004).

### 2.2 Recruitment and sampling

This study focused on the policy, health service, and clinical implications of an NEML in Ireland. Interview participants were identified as those employed or holding a position in a body, group or organisation with a role in prescribing/medicines policy or national therapeutics choices and likely immediately involved in the political and clinical decision-making process surrounding the development and implementation of an NEML. Therefore, a mix of purposive and snowball sampling was used to recruit interest-holders from key sectors/organisations with a role in shaping medicines policy and/or use in Ireland, seeking at least one participant from several of those listed in eBox 1, as well as from other sectors (advocacy, academia/education, prescribers). Given the narrow study focus, the small number of people relevant to the EML policy process in Ireland, and considering other interview studies of Irish policy makers [15], at least one participant from those listed in eBox 1 was considered sufficient in terms of information power [16].

An email invitation was circulated to potential participants identified by REDACTED, which included a participant information leaflet and link to a consent form.

### 2.3 Data Collection

Semi-structured interviews were conducted by REDACTED between May-October 2023. REDACTED had no prior interactions with most interviewees, except two who were current or former colleagues. Interviews were conducted via Microsoft Teams and lasted 30-45 minutes. A topic guide guided interviews (see eBox 2), focusing on the NEML’s purposes and potential NEML development processes. The topic guide was developed by REDACTED and REDACTED with additional recommendations provided by the research team. The topic guide was piloted and refined before data collection commenced.

### 2.4 Data Analysis

Interviews were audio-recorded and initially transcribed using Microsoft Teams. One author (REDACTED) read the transcripts and listened to the audio recordings to correct any inaccuracies. Participants were given the opportunity to review their transcripts and correct any inaccuracies. Participants were also offered the opportunity to view a final draft of the results. One author (REDACTED) conducted thematic analysis using Braun and Clarke’s approach [17–19]. A mix of inductive and deductive thematic analysis was used. REDACTED read and reread the transcripts, and then conducted line-by-line coding, organising meaningful text segments into initial codes and categories. By comparing codes across transcripts, REDACTED then refined and labelled the codes consistently. These were then inductively synthesised into subthemes which were deductively assigned to one of three overarching themes, corresponding to the main components of the topic guide. Additionally, during the transcription process outlined above, the relevant author (MP) developed codes and themes. These were used by the primary coder (REDACTED) to sense check the analysis. To increase the confirmability of the findings, an author (REDACTED) crosschecked the coding structure and themes developed. Themes are reported with illustrative quotes and deidentified participant codes. Analysis was conducted using NVIVO Version 12.

### 2.5 Reflexivity

The researchers involved in the analysis discussed their professional experience, potential biases, and preconceptions about the research area throughout data analysis. Regarding positionality and reflexivity for the research team, three authors were registered pharmacists, two licensed in Ireland (REDACTED and REDACTED) and one in the USA (REDACTED). One study author (REDACTED) had existing relationships with policymakers and other interest-holders. All study authors had familiarity with the NEML literature and intended purposes of an NEML. REDACTED, who conducted interviews, identifies as male, was a postdoctoral researcher when conducting interviews, held a PhD, and had training and experience of qualitative interviewing. REDACTED was employed to conduct interviews and his knowledge of NEMLs at the point of interviews primarily related to information in the protocol.

## 3. Findings

Of fourteen people invited to interview, thirteen were each interviewed once, one person declined interview. Interviewees included individuals employed by some, but not all, bodies with roles in medicines policy/use in eBox 1, as well as practising clinicians with specific prescribing expertise or involvement in relevant non-governmental organisations. Three main themes were developed: 1) the purpose of an NEML, 2) barriers and facilitators for NEML implementation and development and 3) development and implementation processes for an NEML, See Figure 1 for an overview of themes.

**Figure 1.**
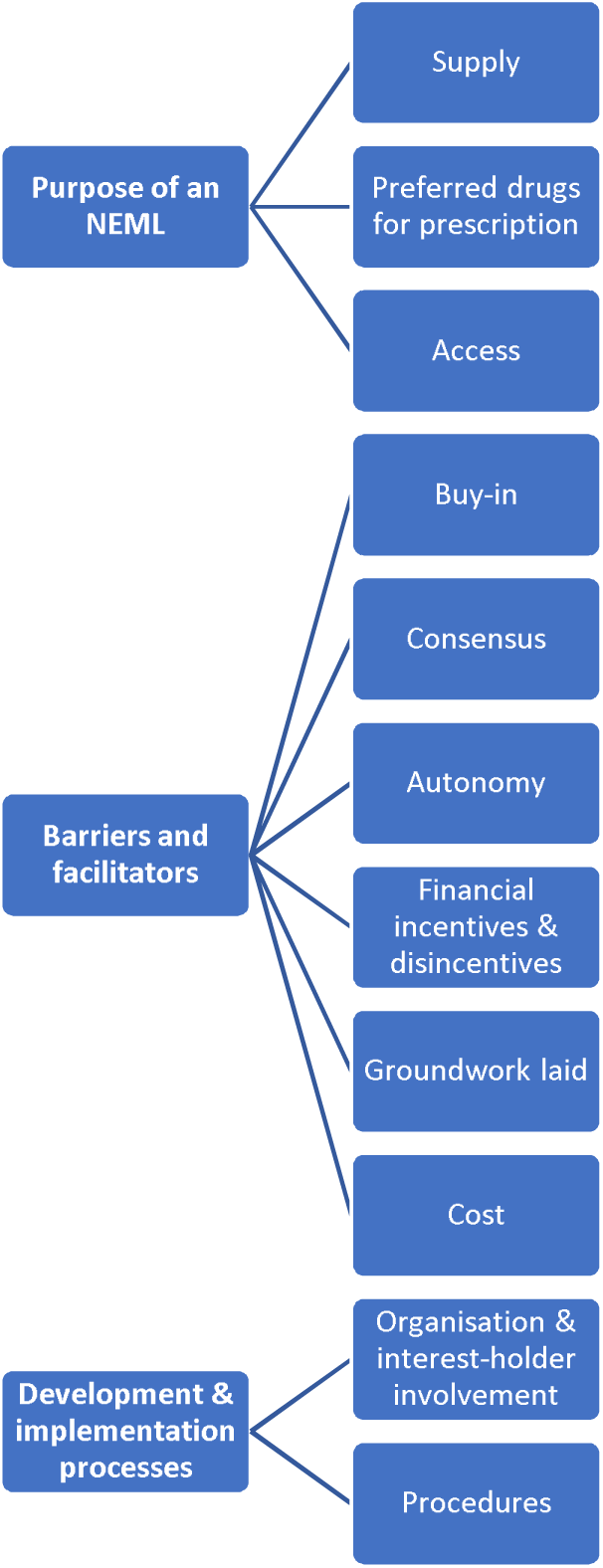
Overview of the themes and subthemes developed.

### 3.1 The purpose of an NEML

Most participants perceived an NEML as meeting “the priority needs of the population” (Interviewee 1 (IV1)) and that an NEML would generally be for common conditions in Ireland. This was conceptualised in different ways. Some referred to a broader list of medicines for an Irish NEML compared to low- and middle-income countries’ (LMICs’) NEMLs, while others primarily thought about the NEML in the context of LMICs. Some gave examples of medicines that should be included. Many discussed lifesaving medicines:

> “[…] what I would see as mission critical, the stuff we cannot do without to deliver basic healthcare, […] for the life threatening” (IV6).

Most participants felt it should be broader, referring to both lifesaving medicines and “life maintenance type of medicines that are used every day [for] common illnesses” (IV5).

Some participants felt that medicines for rare diseases should be included because they “are a matter of life and death” (IV5). However, most thought that rare disease medicines should not be included. Participants thought the emphasis should be on “The priority needs of the population, conditions which are common and affect lots of people. Not to say that drugs for rare conditions wouldn’t be considered essential, but often those medicines might be more expensive, or they might be newer, or there might be less evidence in terms of effectiveness.” (IV1)

#### 3.1.1 Supply

> “An [NEML] ultimately serves the purpose of can we provide these basic medicines to our population, and have we got a sure supply of them?” (IV3)

Participants highlighted the potential role of NEMLs in strengthening supply chains by mapping sources and agreements for NEML medicines and ensuring their robustness. Participants also suggested that it might involve legal requirements to have stockpiles of these medicines (e.g. 1-3 months’ supply). One participant said stockpiling only “delays something” but “can be useful if there’s a short-term problem” (IV11), and suggested examining shortages through two lenses: availability of therapeutic alternatives, and patient impact.

Some participants mentioned an NEML leading to centralised procurement but one participant discussed challenges with that:

> “Centralised procurement. We don’t do that, which makes these kinds of things more difficult to implement because there’s no system at the moment that you can log in and say ‘what level of this antibiotic do we have in Ireland?’” (IV9)

#### 3.1.2 Preferred drugs for prescribing

> Participants also envisioned an Irish NEML as: “A national essential medicines formulary or list: having a system where most prescribers who are prescribing most of the time for common things, prescribe from an NEML.” (IV1)

Some saw it as influencing formularies in specific settings:

> “I can see it influencing a formulary for hospitals. Each individual hospital works as its own entity and controls its own budgets, so there may not be a desire to have a coordinated approach.” (IV11)

Some participants envisioned the NEML involving formulary and supply chain measures existing alongside each other:

> “You want to make sure they are available, […] that they are the most effective drugs available. But carefully balanced with the cost to the health system” (IV2).

#### 3.1.3 Access

Some participants envisioned “a universal list of essential medicines that was going to be made available to everyone, that this would be a more refined list than what is currently available.” (IV1)

However, several participants felt that it should not inform entitlement decisions:

> “I’m not convinced policy wise, [universal access to the NEML] would work here.” (IV6)

Most participants felt that the NEML would fit into the current entitlements system:

> “I would imagine it would run in the same way we run our usual reimbursement schemes […], if you have an ability to pay, you pay, or if [not], then you are provided with those free of charge or with a fee.” (IV3)

### 3.2 Barriers and facilitators

Participants detailed several barriers and facilitators to NEML development and implementation.

#### 3.2.1 Buy-in

Participants discussed the need to “get buy-in” (IV7) for an NEML from several organisations and groups, including the national government health department and the body who run the public health service (Health Service Executive, HSE) (details in eBox 1), as well as prescribers, the government/politicians, patients and the public. To get buy-in from the latter groups, participants discussed the importance of communication and framing (see Table 1, quote 1), as NEML’s could be perceived as cost-cutting measures or efforts to reduce access or prescriber freedoms.

**Table 1.** Key quotes relating to the barriers and facilitators to National Essential Medicines List (NEML) development and implementation.

| Subtheme | Component | Relevant Quote |
| --- | --- | --- |
| Buy-in | Communication | 1. "It's about communicating the message and it's communicating the message to the patient groups, and the clinicians, and the public. And if you don't get that right, it won't work." (IV6) |
| Consensus | How many drugs within a class | 2. "If we use a category of within gastrointestinal and we say we need a certain number of proton pump inhibitors, which is a class within gastrointestinal, are they OK with one or two, you know? Or do they want three [...] And then whether you have multiple ones, but actually define that the essential list must have one of these, or two of these, so, from a supply perspective, would the clinicians involved be OK with actually interchanging between these." (IV3) |
| Autonomy | Preferred drugs | 3. "So what that looked like was, just like the UK, we go with a preferred agent. It's like the Medicines Management Programme, but extended out to more than the number of conditions that there currently is. So, we say you'd have a preferred corticosteroid inhaler, a preferred reliever inhaler, you'd have a preferred antihypertensive. You know different one, for different class of antihypertensive and that would be based on evidence and cost." (IV4) |
| Financial Incentives and | Incentives | 4. "Years ago, there was this scheme [generic medicines initiative*] where the government actually paid GPs to prescribe within a certain formulary to |
| Disincentives |  | change things over, and then they could get a grant from it, to do up their practice.” (IV4) |
| Groundwork laid | Unified budgets could facilitate NEML | 5. “We’ve got a finite amount of resources, we’ve one pot of money it’s split in different places at the moment. Hopefully their reorganisation with Sláintecare, where the different heads of those RICOs [Regional Integrated Care Organisations] are responsible for primary and secondary care, will make things a bit better because it doesn’t become then my precious and your precious” (IV13) |
| Cost | NEML increasing purchasing power | 6. “I suppose the main purpose would be to reduce unwanted variation, to improve the quality of prescribing, to reduce costs. And then I suppose on a larger scale, if for example, a more refined, or smaller set of medicines were available, under some sort of payment scheme, that if everyone for example, was prescribing the same ACE inhibitor, or PPI, or whatever it is. That would give some sort of buying power, and the government may be able to negotiate a better cost with the manufacturer, or the drug company that owns that particular medicine.” (IV1) |
| Cost | NEML would be leverage to increase prices | 7. “Any company that wants to put that product on the market, is it going to say this is an essential medicine, I need a premium in order to get this product to the market.” (IV11) |

\*The generic medicines initiative is no longer an active scheme

#### 3.2.2 Consensus

Participants generally thought that getting “consensus” (IV3) on what medicines to include in an NEML would be challenging. They thought that clinicians and patient groups might push excessively for medicines from their clinical area:

> “Different organisations look after different things; they’re going to be concerned about their patch.” (IV13)

One participant discussed exempt medicinal products (i.e. unlicensed medicines) and their place on an NEML:

> “But what about dexamethasone? It’s not authorised for [COVID-19], but it’s absolutely vital and that leads into another conversation: what about this? What about that? So, it becomes tricky” (IV11)

That participant elaborated that this might encourage processes for authorising some off-label usages and another participant mentioned that an NEML “could encourage research into those conditions” (IV2).

Participants were also concerned about consensus regarding the threshold for inclusion:

> “what’s basic healthcare in Europe and Ireland, […] it’s diverging. Our expectations around our healthcare are quite high.” (IV6)

One participant suggested that this could become problematic:

> “What it defines is a lower threshold of what we basically need. If it starts to be used as an indicator of where we’re wanting to place ourselves in terms of basic care, then it becomes something that could be problematic.” (IV3)

Consensus on how many drugs to include within a class was also a concern (see Table 1, quote 2).

#### 3.2.3 Autonomy

Regarding a national formulary, some participants cautioned against rules restricting the medicines clinicians can prescribe. Participants spoke of clinicians distrusting organisations like the HSE:

> “Clinicians can be hesitant to messaging coming directly from say the HSE because ‘they’re only thinking about the money.’” (IV2)

Some participants proposed a “stick and carrot approach” (IV12), with the suggestion that a version of the HSE Medicines Management Programme’s (see eBox 1) preferred drug initiative could be an option (see Table 1, quote 3).

#### 3.2.4 Financial incentives and disincentives

Participants said prescribers would feel disincentivised if the savings resulting from NEML adherence went into general government funds or were used for areas perceived as wasteful:

> “There needs to be a concrete ‘we’re going to fund X, Y, & Z,’ there needs to be carrots.” (IV13)

One participant described a previous system of financial incentives no longer in place (see Table 1, quote 4).

#### 3.2.5 Groundwork laid

Several participants discussed existing work that could inform an Irish NEML. This primarily surrounded a “critical medicines list” (IV11) that was developed because of supply chain issues arising from the COVID-19 pandemic and the UK leaving the EU. Participants also discussed how NEMLs had been developed in other European countries. Participants mentioned other EU initiatives related to NEMLs that could inform an Irish NEML, including the WHO Model List of Essential Medicines, Ireland’s national reimbursement list, and the Heads of Medicines Agencies/European Medicines Agency Task Force on Availability of Authorised Medicines for Human and Veterinary Use.

Participants discussed how Ireland’s health reform policy, Sláintecare, could indirectly support an NEML. Sláintecare was seen as a significant shift in healthcare policy in Ireland within which an NEML could be incorporated. Participants also noted NEML-related changes introduced under Sláintecare including “the community hospital systems coming together as one” (IV10) and budget unification (see Table 1, quote 5).

#### 3.2.6 Cost

Cost was considered both a barrier and facilitator. As a barrier, time and resources would be needed for people/organisations to develop and maintain the NEML. Some felt that the NEML might save money by increasing purchasing power and reducing use of costlier and less effective medicines (see Table 1, quote 6). Others cautioned that increased purchasing power could reduce the number of suppliers due to a reduced market for their medicines which could influence shortages and prices. It was cautioned that NEML medicine prices could increase as companies might use a medicine’s NEML status as leverage (see Table 1, quote 7).

Some participants said that for actors such as wholesalers, there “would have to be a cost associated with” strengthening supply chains but that this may be justified:

> “In terms of a policy decision you have the reason for why you need to spend this money” (IV9)

### 3.3 Development and Implementation Processes

#### 3.3.1 Organisation and interest-holder involvement

The main organisations that participants proposed should develop and maintain the NEML were the national government health department, the HSE or the national medicines and devices regulator (Health Products Regulatory Authority, HPRA) (see eBox 1) or a combination of those organisations (see relevant quotes for these interest-holders and others in Table 2).

**Table 2.** Key quotes relating to organisation and interest-holder involvement in a National Essential Medicines List (NEML).

| Components | Organisation/groups | Quote |
| --- | --- | --- |
| Clinical | National clinical programmes | "Obviously there's all the different national clinical programs from the HSE that might have an input into all of this. So they should be consulted." (IV8) |
| Regulator | Health Products | "There's the medicine regulator because we |
|  | Regulatory Authority | regulate the authorization of medicines, we regulate the manufacturers and the wholesalers and lots of other things in between. And then in Ireland, as in lots of European countries, we coordinate the response to shortages.” (IV11) |
| Public/Patients |  | “It would certainly have a lot more buy in if you did have that perspective of a patient there” (IV3) |
| Academia |  | “I do think academics [...] because it brings a bit a bit more of a systematic approach sometimes.” (IV11) |
| Governmental organisations | National government health department and Health Service Executive (HSE) | “But obviously you’d need the strategic arm and you need your operational arm. So you would need your Department of Health, and then you would need your HSE.” (IV13) |
| Commercial bodies | Wholesalers | “The wholesalers, I would consider important because they're the supply chain backbone, they get the stuff out to the patients, the healthcare professions” (IV11) |
|  | Pharmaceutical companies | “So, you're talking about the manufacturers, the owners of the products, the marketing authorization holders, you're talking about the distributors.” (DS) |
| Multiple |  | “So patients, professionals, academia, government agencies, people like the regulator who are going to be securing supply.” (IV6) |

There were several technical, clinical, policy and other organisations/groups who participants thought should provide input on an NEML. Clinical groups included the national clinical programmes, national clinical leads, and clinicians with relevant specialist knowledge. Given the importance placed on cost-effectiveness, the National Centre for Pharmacoeconomics was stressed as important, while others said that only reimbursed medicines should be included. For the HSE, several participants suggested input from specific departments or groups such as the acute hospital drugs management programme.

From the commercial side, participants said there should be input from pharmaceutical companies and wholesalers. However, participants stated that pharmaceutical companies’ influence should be limited: “they’re not meant to be the decision-maker and we blur that a bit here in [Ireland].” (IV6)

Most participants said that patients, patient groups and the public should have input.

#### 3.3.2 Procedures

Participants discussed a “steering group” overseeing the NEML development process, with “subgroups that might work through what the essential medicines would be” (IV5)

Participants also emphasised that it is “important from the start, [to be clear about] the purpose of this list” (IV7). Participants suggested that an NEML should be updated periodically, but that “developments in the healthcare system […] might trigger earlier reviews.” (IV7). One participant recommended “an evaluation alongside [the NEML rollout].” (IV1)

## 4. Discussion

Thirteen Irish health system interest-holders were interviewed about a potential NEML in Ireland. Participants described the purpose of an NEML as meeting the priority needs of the population. They also outlined possible roles for an NEML in ensuring adequate supplies and as a national formulary. Views differed on whether an NEML should involve reduced costs to patients for access. Several barriers (e.g. consensus) and facilitators (e.g. buy-in) to developing and implementing an NEML were outlined. Regarding practicalities, participants outlined that the national government health department, the HSE, and/or the HPRA should be responsible for the NEML.

Participants’ views on an NEML aligned with the WHO’s: that the list includes medicines that are “available, affordable” and “effectively and safely treat the priority healthcare needs of the population” [20]. Affordability is important in the WHO Model List of Essential Medicines, though “authors close to” its processes [21] have advocated for greater focus on affordability [22]. The WHO Model List has newer expensive medicines that are considered unaffordable with no low-cost alternative [23], which undermines potential cost-savings associated with NEMLs. This raises questions about what “essential” means [24], and whether the WHO List is primarily for LMICs [21, 23, 25]. Our participants also discussed these issues. Some authors have called for the WHO to articulate the target audience for their Model List [26], though it has been suggested that this ambiguity is a political strategy, to facilitate several interests [21]. The original 1977 WHO Model List focussed on LMICs and affordable medicines [26]. Procedures were revised in 2001 and price and patent were no longer considered possible reasons for exclusion [26], making the WHO List more relevant to high-income countries (HICs) [26]. Twenty-one of 79 HICs had NEMLs in 2017 [27], and this is increasing [28]. Given that most HICs’ NEMLs are similar [27], it is feasible for those NEMLs to inform an Irish NEML, as suggested by our participants and the WHO [6].

Some have expressed concern about pharmaceutical industry influence on a list [24]. Some of our participants also warned of this. Pharmaceutical industry interests have influenced WHO EML development processes [21]. This relates to opposing views about whether the WHO Model List reduces or increases prices [21]. Our participants echoed such views.

The Swedish “Wise List”, a HIC NEML, has 200 “core medicines” for treating common diseases in primary care and secondary care and “100 complementary medicines for common diseases in specialised care” [29]. According to a 15-year retrospective study, this NEML has effectively increased the prescribing of cost-effective medicines [29]. High adherence to the list has been ascribed to 1) transparent development processed with conflict of interest management and 2) the communication strategy [29]. A qualitative study of interest-holders in Canada, also emphasised the importance of transparency and communications strategy [24]. Communication was emphasised as very important by our participants. Questions have been raised about the transparency of WHO processes [21], despite WHO guidance emphasising this.

In Sweden, there are NEML financial incentives and disincentives; NEML medicines are free of charge in hospitals and 50% of the cost of non-NEML medicines are funded by the hospitals, while local financial incentives apply in primary care in Stockholm [29]. Our participants also discussed NEML financial incentives and disincentives. Along with the historical example cited by a participant [30], this has a recent precedent in Ireland; the Medicines Management Programme introduced a “best-value biological” initiative whereby hospital teams receive €500 if they initiate a patient on a biosimilar [31, 32]. Also, there are financial disincentives, as biologics with biosimilars available are not reimbursed [33]. Biosimilar prescribing has increased since both measures were introduced [34].

In 2020, the US Food and Drug Administration issued an executive order to “identify the list of essential medicines, […] that are medically necessary to have available at all times” [35] with a focus on strengthening “domestic supply chains” [35]. Our participants spoke in general terms about supply chains, with some reference to European initiatives. There have been further recent EU developments such as the Critical Medicines Act which was proposed in 2025 and aims “to improve the availability, supply and production of critical medicines within the EU” [36]. The US document also mentions affordability for veterans [35]. In Canada, a comprehensive approach was considered; a 2019 government report called for state funding of a “list of essential medicines” [37]. There were contrary views among our participants about whether an NEML in Ireland should lead to increased patient entitlements to medicines, with most suggesting that it should not. Increased entitlements can lead to increased access and adherence to essential medicines, but increased direct costs to the state [38], though potentially lower costs overall [9].

Participants discussed how Ireland’s health reform policy, Sláintecare [39], supports an NEML. One recommendation of the first 2017 Sláintecare report, was to examine “models in use internationally to identify best practice in medicines” [40]. NEML development could be considered in the next Sláintecare action plan.

Some processes suggested by our participants align with WHO NEML guidance [6]. The WHO recommend the national government health department establish an expert committee to select medicines with support from a “technical and administrative secretariat”, and that the approach should align with national medicine selection and reimbursement processes [6]. The only differences between WHO guidance and the views of our participants were that participants suggested input from technical agencies rather than a technical secretariat, and involvement of subcommittees for disease areas. Others have also suggested greater focus on disease areas in the WHO Model List [26]. The WHO List is updated every two years, and the WHO recommend NEMLs be reviewed and updated at least every two years after WHO List updates [6]. The WHO also suggest triggers for updates, e.g. if the NEML and WHO List start diverging [6]. These processes were similar to those suggested by our participants.

Any individual, organisation, company, or country can apply to change the WHO Model List [20], and comments can be submitted to the WHO secretariat on these applications, with applications and comments published. These processes have been criticised as leading to a narrowly focused applicant-led process [26]. Though the WHO secretariat solicits applications if gaps on the EML are identified [26]. Such processes were not suggested by our participants, though they did suggest wide input (e.g. industry, patient groups) on NEML development. The WHO Expert Committee on Selection and Use of Essential Medicines is required to have “strong clinical and technical expertise in medicine evaluation and clinical use” [20]. It has been suggested that funding is lacking for the Model List’s processes [21]. Funding was also considered a barrier by our participants.

### 4.1 Strengths and Limitations

A strength is that semi-structured interviews facilitated a flexible approach and the exploration of several areas. This is also the first study on NEMLs in Ireland, with limited qualitative exploration of NEMLs in HICs. Regarding limitations, coding by two people independently could have increased data analysis reliability, however, our approach facilitated reflexivity. Interviews with interest-holders of this nature can lead to limitations such as omission and social desirability bias [41]. Lack of input from patients, the public, and the pharmaceutical industry is also a limitation [21]. Finally, the low number of participants is a potential limitation; however, we considered the study to have sufficient information power given its narrow focus [16].

### 4.2 Conclusion

Although more evidence is needed on the impact of NEMLs on medications costs and ultimately the health of individuals and population, interest-holders in Ireland see potential benefits of an NEML in Ireland. Participants envisioned an NEML that meets the priority needs of the population, ensures adequate supplies and acts as a national formulary.

## Supporting information

eBox 1

## Data Availability

Due to the potential for participants to be identified as the nature of the data prevents full anonymisation, transcripts are not available.

## Acknowledgements

We would like to acknowledge all those who took part in this study, we are grateful for their time and expertise.

