## Supplementary material for "An essential medicines list in Ireland: A qualitative interview study of interest-holders": eBox 1

By REDACTED

### eTable 1. Consolidated criteria for reporting qualitative studies (COREQ): 32-item checklist

| **No. Item** | **Guide questions/description** | **Reported on Page #** |
| --- | --- | --- |
| **Domain 1: Research team and reﬂexivity** |  |  |
| *Personal Characteristics* |  |  |
| 1. Inter viewer/facilitator | Which author/s conducted the inter view or focus group? | 12 |
| 2. Credentials | What were the researcher’s credentials? E.g. PhD, MD | 12 |
| 3. Occupation | What was their occupation at the time of the study? | 12 |
| 4. Gender | Was the researcher male or female? | 12 |
| 5. Experience and training | What experience or training did the researcher have? | 12 |
| *Relationship with participants* |  |  |
| 6. Relationship established | Was a relationship established prior to study commencement? | 12 |
| 7. Participant knowledge of the interviewer | What did the participants know about the researcher? e.g. personal goals, reasons for doing the research | 12 |
| 8. Interviewer characteristics | What characteristics were reported about the inter viewer/facilitator? e.g. Bias, assumptions, reasons and interests in the research topic | 12-13 |
| **Domain 2: study design** |  |  |
| *Theoretical framework* |  |  |
| 9. Methodological orientation and Theory | What methodological orientation was stated to underpin the study? e.g. grounded theory, discourse analysis, ethnography, phenomenology, content analysis | 12 |
| *Participant selection* |  |  |
| 10. Sampling | How were participants selected? e.g. purposive, convenience, consecutive, snowball | 11 |
| 11. Method of approach | How were participants approached? e.g. face-to-face, telephone, mail, email | 11 |
| 12. Sample size | How many participants were in the study? | 14 |
| 13. Non-participation | How many people refused to participate or dropped out? Reasons? | 14 |
| *Setting* |  |  |
| 14. Setting of data collection | Where was the data collected? e.g. home, clinic, workplace | 11-12 |
| 15. Presence of non-participants | Was anyone else present besides the participants and researchers? | N/A |
| 16. Description of sample | What are the important characteristics of the sample? e.g. demographic data, date | N/A |
| *Data collection* |  |  |
| 17. Interview guide | Were questions, prompts, guides provided by the authors? Was it pilot tested? | 11 |
| 18. Repeat interviews | Were repeat inter views carried out? If yes, how many? | N/A |
| 19. Audio/visual recording | Did the research use audio or visual recording to collect the data? | 12 |
| 20. Field notes | Were ﬁeld notes made during and/or after the inter view or focus group? | N/A |
| 21. Duration | What was the duration of the inter views or focus group? | 11 |
| 22. Data saturation | Was data saturation discussed? | N/A |
| 23. Transcripts returned | Were transcripts returned to participants for comment and/or correction? | N/A |
| **Domain 3: analysis and ﬁndings** |  |  |
| *Data analysis* |  |  |
| 24. Number of data coders | How many data coders coded the data? | 12 |
| 25. Description of the coding tree | Did authors provide a description of the coding tree? | 12 |
| 26. Derivation of themes | Were themes identiﬁed in advance or derived from the data? | 12 |
| 27. Software | What software, if applicable, was used to manage the data? | 12 |
| 28. Participant checking | Did participants provide feedback on the ﬁndings? | N/A |
| *Reporting* |  |  |
| 29. Quotations presented | Were participant quotations presented to illustrate the themes/ﬁndings? Was each quotation identiﬁed? e.g. participant number | 15-24 |
| 30. Data and ﬁndings consistent | Was there consistency between the data presented and the ﬁndings? | 15-24 |
| 31. Clarity of major themes | Were major themes clearly presented in the ﬁndings? | 14 |
| 32. Clarity of minor themes | Is there a description of diverse cases or discussion of minor themes? | 14 |

### eBox 1. Overview of organisations and sectors involved in medicines policy and use in Ireland

**Health Service Executive (HSE):** Responsible for running all of the public health services in Ireland.**^1^**

**Department of Health:** Strategic responsibility for health and personal social services in Ireland.^2^

**Health Products Regulatory Authority (HPRA):** The national regulator of medicines and devices.^3^

**National Clinical Programmes:** Multi-disciplinary teams designed around population groups, medical specialties, or specific diseases, to develop healthcare services in Ireland, for example through development of clinical pathways, models of care and clinical guidelines.^4^

**National Centre for Pharmacoeconomics:** A health technology assessment agency with responsibility for evaluating medicines to be used by the healthcare system in Ireland.^5^

**HSE Medicines Management Programme:** A multi-disciplinary team who lead nationally on medicines management processes, access to medicines and medicine expenditure.^6^

**RICOs (Regional Integrated Care Organisations) or Health Regions:** Six organisations responsible for planning and delivering health and social care services in their region of Ireland.^7^

**HSE Acute hospital drugs management programme:** Provides strategic oversight of medicine use in hospitals, supporting safe, efficient and cost-effective medicine use. It is one of three elements of the Irish Access and Integration Drug Management Programme.^8^

**National Clinical Effectiveness Committee:** A national body that provides a framework for the endorsement of clinical guidelines at a national level.^9^

**HSE Antimicrobial Resistance & Infection Control (AMRIC)**: An action plan and governance groups with the goal of coordinating a cross-sectoral approach to antimicrobial resistance.^10^

**Professional Bodies**: There are several professional bodies (e.g. Irish College of General Practitioners) in Ireland that play a variety of roles (e.g. medical education, training programmes). Professional bodies often provide input on clinical guideline development.

### eBox 2. Topic Guide

**Stakeholder perspective on the potential for an essential medicines list in Ireland**

Pre Q – Do you need anything clarified from the information leaflet?

Introduction:

So, just to reiterate from the information leaflet, your responses will be pseudonymised with any potentially identifying details removed. Your participation is voluntary and you may choose to stop the interview at any time. Have you any questions?

Can I check are you happy for me to start recording now?

**Perspectives and Knowledge on EML**

I’d like to start by asking you a few questions about your perspective on essential medicines lists and how an EML could be used in Ireland.

1. What do you understand by the terms “essential medicines” and an “essential medicines list”?

**Definition:** An Essential Medicines List, as defined by the WHO, is a list of medicines which meet the priority health needs of a population.

**I. Potential Applications of EML in Ireland**

2. What is your perception of how national essential medicines lists are used in a global context?

3. What purposes could you foresee a national essential medicines list serving in Ireland?

a. How might a national essential medicines list impact medication access costs?
b. How might a national essential medicines list impact clinician practice?
c. How might a national essential medicines list impact medication access and shortages?
d. How might a national essential medicines list impact patient safety?

4. How do you see an Irish list differing from how you’ve described it being used in a global context

**II. Development and Implementation**

4. What would be the enablers and barriers of developing and implementing a national essential medicines list?
   a. These can be in general, or relating to specific purposes a list could have, such as procurement or medication entitlements.
5. What factors or criteria do you believe should be considered when deciding which medicines should be included in a national essential medicines list for Ireland?

Probe:

- Costs?
- Disease prevalence (how many people have each disease)?
- Specific patient groups or rare conditions factors?

6. Who or what organisation within the health system should have responsibility for	developing and updating such a list?
   a. Which stakeholders do you think will be most important in determining whether an essential medicines list would be implemented? Why?

7. How do you believe decisions around developing and updating an EML should be made from a process perspective?
   a. For example, the process could be closed to a small selection committee, or a process that is open and transparent that accepts inputs from the public.
8. what stakeholders do you think should be involved in developing and updating an EML?
   a. These can be representatives (technical, implementation or political) from various bodies/organisations within the health system, pharmacists, specialists and general practitioners, researchers, the public, patients, etc.

**III.NEML Fit with Existing Structure/Processes/Policies/Political Climate/Context**

9. How would a national essential medicines list fit alongside existing systems and policies that influence medicines use?
   a. E.g., medication entitlements, guideline recommendations
10. How would a national essential medicines list fit within ongoing health reform?
11. What are the political barriers to implementing an EML?

(probe: what political factors might contribute to the development of a national essential medicines list?)

12. What are your perspectives on the general mood surrounding health reform in Ireland?
a. Probe: How would an essential medicines list contribute to the mood surrounding health reform in Ireland?

**IV. Conclusion**

13. To finish, is there anything else you would like to add that we haven’t discussed so far?

Thank you for your time and for taking part in our research study. We would like to give you the opportunity to review the transcript of your interview today. Are you happy for us to contact you again when this is available for review?

We are also interested to hear if you have a colleague or contact who you think it would be helpful for us to interview. If so, would you be happy to pass on an invitation to them on our behalf?
